# Exploring technology access, knowledge, and preferences for detection, monitoring, and support of cognitive health in rural communities

**DOI:** 10.1101/2025.06.11.25329469

**Authors:** Michael J. McCarthy, Megan C. McCoy, Travis Anderson, Rasheera Dopson, Margarita Martinez, Thomasina J. Seaton, Duy Anh Duong, Eric S. Cerino

## Abstract

Approximately 25% of adults in the United States aged 60 years and older experience Mild Cognitive Impairment (MCI) or Subjective Cognitive Decline (SCD) which may be precursors to Alzheimer’s Disease. Families living in rural areas and contending with MCI/SCD face unique challenges due to geographic isolation, lack of specialty healthcare providers, and limited access to information and resources that support brain health. Thus, digital health technologies hold promise for improving cognitive health in rural communities. This study explored the access, knowledge, and preferences of rural individuals with MCI/SCD and their family members for detection, monitoring, and support of cognitive health. A convergent mixed methods design was used with semi-structured interviews informed by the Senior Technology Acceptance Model, paired with descriptive survey data from N=20 individuals with MCI/SCD and their family members. The majority of participants had access to and knowledge of technologies to support cognitive health, although preferences for detection, monitoring, and support differed between individuals with MCI/SCD and family members. Participants described aspects of technology that they valued (e.g., to organize and track tasks, to “exercise” one’s brain, to communicate with healthcare providers, to promote safety of the person with MCI/SCD), as well as concerns that they had (e.g., “generational” barriers to adoption and use, financial cost, privacy and security, lack of human interaction). Findings from this study will directly inform the development of supportive technologies for meeting the needs of rural families experiencing MCI/SCD.

**Author Summary:** Rural “memory care deserts” are characterized by lack of specialty healthcare providers and lack of basic information and resources to support brain health. Although telehealth may be an option in such places, there are a variety of barriers that impede widespread implementation including lack of infrastructure and financial resources, limited high-speed internet, interstate provider and billing issues, and general mistrust among users of technology, as well as non-rural healthcare providers. Thus, digital health technologies that are inexpensive, user-friendly, sustainable, and delivered within community settings such as senior centers may hold promise for supporting brain health in rural communities. To successfully develop these technologies, however, we must have a deep understanding of the perspectives of potential users so that we may leverage what they value in technology (e.g., ability to organize and track tasks, to “exercise” the brain, to communicate with providers), as well as address the concerns that they have (e.g., cost, privacy and security, lack of human interaction). This mixed-methods study (qualitative interviews, survey data) explores these issues and provides information to directly inform development of brain health resources for rural memory care deserts.

## Introduction

Approximately 25% of adults in the United States aged 60 years and older experience Mild Cognitive Impairment (MCI) or Subjective Cognitive Decline (SCD) (1,2). MCI refers to an intermediary stage between normative cognitive aging and dementia characterized by more memory and thinking challenges than age- and education-level peers (3,4). SCD refers to self-reported awareness and concerns with declines in one’s cognition (5). Individuals with MCI or SCD (henceforth MCI/SCD) exhibit a variety of noticeable symptoms including forgetfulness, inability to maintain their train of thought or track conversations, difficulty with wordfinding, decision-making, and navigating familiar places, and generally poor judgement (6). These symptoms are distressing for the individual affected as well as their family members who may experience uncertainty, social problems, sleep disruptions, stress, burden, and anxiety (7,8). MCI/SCD may also be important precursors to Alzheimer’s Disease (AD) (9) which is extremely costly, both in economic as well as human terms [AAA, 2025].

Families living in rural areas and contending with cognitive problems are often isolated (10), underdiagnosed (11), have few resources (12), experience geographic barriers to care (10), and lack basic access to cognitive information and services (13,14), including community-based preventative “brain health” education and resources. These “memory care deserts” can be difficult to navigate for families and, as a consequence, MCI/SCD symptoms can go untreated and unchecked, potentially developing into clinical dementia (15). Studies suggest that, compared with urban individuals, rural individuals with dementia experience more days in nursing homes and significantly higher mortality (16,17).

Although telehealth approaches to detecting and treating MCI/SCD and dementia have gained prominence in recent decades (18), rural areas face barriers to adoption of telehealth such as limited healthcare infrastructure, limited access to high-speed internet, limited financial resources necessary for installation and sustainability of telehealth technologies, challenges with interstate provider licensing and billing, and general mistrust among users of technology as well as non-rural healthcare providers (19–22). As such, digital health technologies that are inexpensive, user-friendly, sustainable, and delivered within community settings may hold promise for addressing these limitations and, thus, improving cognitive health in underserved memory care deserts (23,24). However, in order to successfully develop and implement these technologies, we must thoroughly understand the perspectives of potential users.

The purpose of this study is to explore the access, knowledge, and preferences of rural individuals with MCI/SCD and their family members for detection, monitoring, and support of cognitive health. Findings will inform the development of supportive technologies for meeting the needs of rural families experiencing MCI/SCD.

## Methods

This study used a convergent mixed-methods design (25), with semi-structured interviews with 10 rural individuals with MCI/SCD and their 10 family members (N=20) recruited through senior centers in Northern Arizona. A detailed description of the study protocol (i.e., Northern Arizona Memory Study) has been published previously (26). Participants were included if they: 1) Lived in the community (i.e., not a nursing home); 2) Could identify a family member who was willing to participate with them; 3) Were capable of using technology, with coaching and support; 4) Were able to communicate in English; and, 5) Were experiencing MCI (based upon Montreal Cognitive Assessment Blind [MoCA Blind] scores ≤18; (27) and/or SCD (based upon adapted items from the Einstein Aging Study [EAS] Health Self-Assessment; (28,29). For MCI, the MoCA Blind was selected due to its capacity for both in-person and remote telephone administration. For SCD, participants answered items on perceived declines in cognition (memory, thinking) and whether they were concerned about these changes, as well as a question on whether they have discussed their memory or thinking challenges with a healthcare provider. SCD was operationalized as self-reported presence of declines in memory or thinking and/or concerns in these domains. Individuals were excluded if they were: 1) Under the age of 18; 2) Pregnant; 3) Had untreated severe mental illness, alcohol/drug abuse, or suicidality; or, 4) Had another neurological condition that significantly impacted function.

Qualitative interviews were paired with a survey capturing participant demographics, subjective health, cognitive status, and information about current access to and use of technology (computer, tablet or iPad, internet or Wi-Fi, Zoom, Skype, Webex, or Google Meet video call, Android smart phone with internet, iPhone with internet), as well as perceived usefulness and comfort with these technologies for MCI/SCD detection, monitoring, and support (Roque et al., under review). Technology items were rated on Likert-type scales from *Strongly Disagree/Extremely Uncomfortable* (*1*) to *Strongly Agree/ Extremely Comfortable* (*5*). A modified version of the *Senior Technology Acceptance Model* (STAM; (30)) informed the qualitative interviews and analysis of participant technology preferences, using the software Dedoose version 10.0.25 (31). Interviews were conducted until saturation was achieved.

Interviews were transcribed by a professional transcription service, spot-checked for accuracy by a graduate research assistant, and then coded by two other graduate research assistants, with a fourth member of the research team overseeing and serving as auditor for the process. Two additional team members provided ongoing feedback as transcripts were coded and key themes were identified. The first author of this paper summarized findings narratively. This summary was reviewed by the other authors to ensure comprehensive and accurate interpretation of data, as well as to promote the credibility, transferability, dependably, and confirmability of qualitative findings (32). Descriptive statistics in SAS 9.4 (33) were used to analyze survey data in order to complement understanding gained through qualitative interviews and analyses.

Participants received Informed Consent and a $50 gift card in appreciation for their time. Research activities were approved by the Institutional Review Board at Northern Arizona University.

## Results

### Sample characteristics

On average, individuals with MCI/SCD were 72 years old (range 32-91) and family members were 58 years old (range 34-75). Half of participants were spousal dyads with the other dyad types including adult children/parents (four dyads), siblings (one dyad), and friends (one dyad). Thirteen of twenty study participants were female (six MCI/SCD, seven family members).

Eighteen of 20 participants indicated their race/ethnicity as “white” (eight MCI/SCD, ten family members). Most participants indicated being financially “comfortable” (five MCI/SCD, six family members), with others indicating having “just enough to pay for living expenses” (four MCI/SCD, two family members) or “not having enough to pay for living expenses” (one MCI/SCD, two family members). Most participants indicated that their subjective health was “excellent” or “very good” (eight MCI/SCD, four family members), with the remainder indicating that their health was “good” (zero MCI/SCD, four family members) or “fair” (two MCI/SCD, two family members). One-hundred percent of participants with MCI also indicated SCD, with seven participants falling at or below the “mild cognitive impairment” range based on the MOCA-Blind (27). (Table 1)

**Table 1.** Characteristics of Study Participants.

| Variable | Individuals with<br>MCI/SCD<br>(n=10) | Family<br>Member<br>(n=10) | Total (N=20) |
| --- | --- | --- | --- |
| Age in years, Mean (range) | 72 (32-91) | 58 (34-75) | 65 (32-91) |
| Relationship |  |  |  |
| Spousal | 5 | 5 | 10 (50.0) |
| Other (e.g., adult child/parent, sibling, friend) | 5 | 5 | 10 (50.0) |
| Sex, n (% of total) |  |  |  |
| Female | 6 | 7 | 13 (65.0) |
| Male | 4 | 3 | 7 (35.0) |
| Race, n (% of total) |  |  |  |
| White | 8 | 10 | 18 (90.0) |
| Other | 2 | 0 | 2 (10.0) |
| Financial status, n (% of total) |  |  |  |
| Comfortable | 5 | 6 | 11 (55.0) |
| Just enough to pay for living expenses | 4 | 2 | 6 (30.0) |
| Not able to pay for living expenses | 1 | 2 | 3 (15.0) |
| Subjective health, n (% of total) |  |  |  |
| Excellent | 4 | 2 | 6 (30.0) |
| Very good | 4 | 2 | 6 (30.0) |
| Good | 0 | 4 | 4 (20.0) |
| Fair | 2 | 2 | 4 (20.0) |
| Poor | 0 | 0 | 0 (0.0) |
| Cognitive status, n (% of total) |  |  |  |
| Subjective cognitive decline* | 10 | n/a | 10 (50.0) |
| Mild cognitive impairment** (at or below) | 7 | n/a | 7 (35.0) |

*Note.* \*Subjective cognitive decline based upon adapted items from the Einstein Aging Study (EAS) Health Self-Assessment. \*\*Mild cognitive impairment based upon Montreal Cognitive Assessment Blind (MoCA Blind) scores $\leq 18$ .

### Survey findings

Survey data indicated that the majority of the sample had adequate *access to* and *knowledge of* technologies for detection, monitoring, and support. The least endorsed items were access to tablets or iPads (60% of participants) and access to iPhones (45% of participants). 100% of participants reported having access to and being knowledgeable about the internet/Wi-Fi.

Among technology options evaluated for MCI/SCD *detection and monitoring*, persons with MCI/SCD preferred written questionnaires (mean=3.3; median=3.5; range 2-4), whereas family members preferred interactive websites with live professional coaching (mean=3.4; median=4; range=1-4). For *receiving support*, persons with MCI/SCD favored interactive websites with live professional coaching (mean=3.4; median=3.5; range=2-4) and had generally low opinions about the usefulness of phone calls or text messages from peers (mean=2.5/2.1; median=2; range=1-4). Family members had similar opinions about all options for receiving support (i.e., phone calls or text messages from live professional coaches, phone calls or text messages from peers, interactive websites or smartphone apps; mean=2.7-3.3; median=3; range=1-4). However, they did not favor written materials for receiving support (mean=2.5; median=2.5; range=1-4).

Participants were generally comfortable with other technologies for better understanding MCI/SCD including activity monitoring data (mean=3.3/3.1; median=3.5/3; range=2-4), heart-rate monitoring data (mean=3.4/3.2; median=4/3; range=2-4), bed-based sleep sensor data (mean=3.2/3.1; median=4/3; range=1-4), and geolocation (GPS) data (mean=2.9/3.3; median=3.5/3; range=0-4). (Table 2).

**Table 2.**
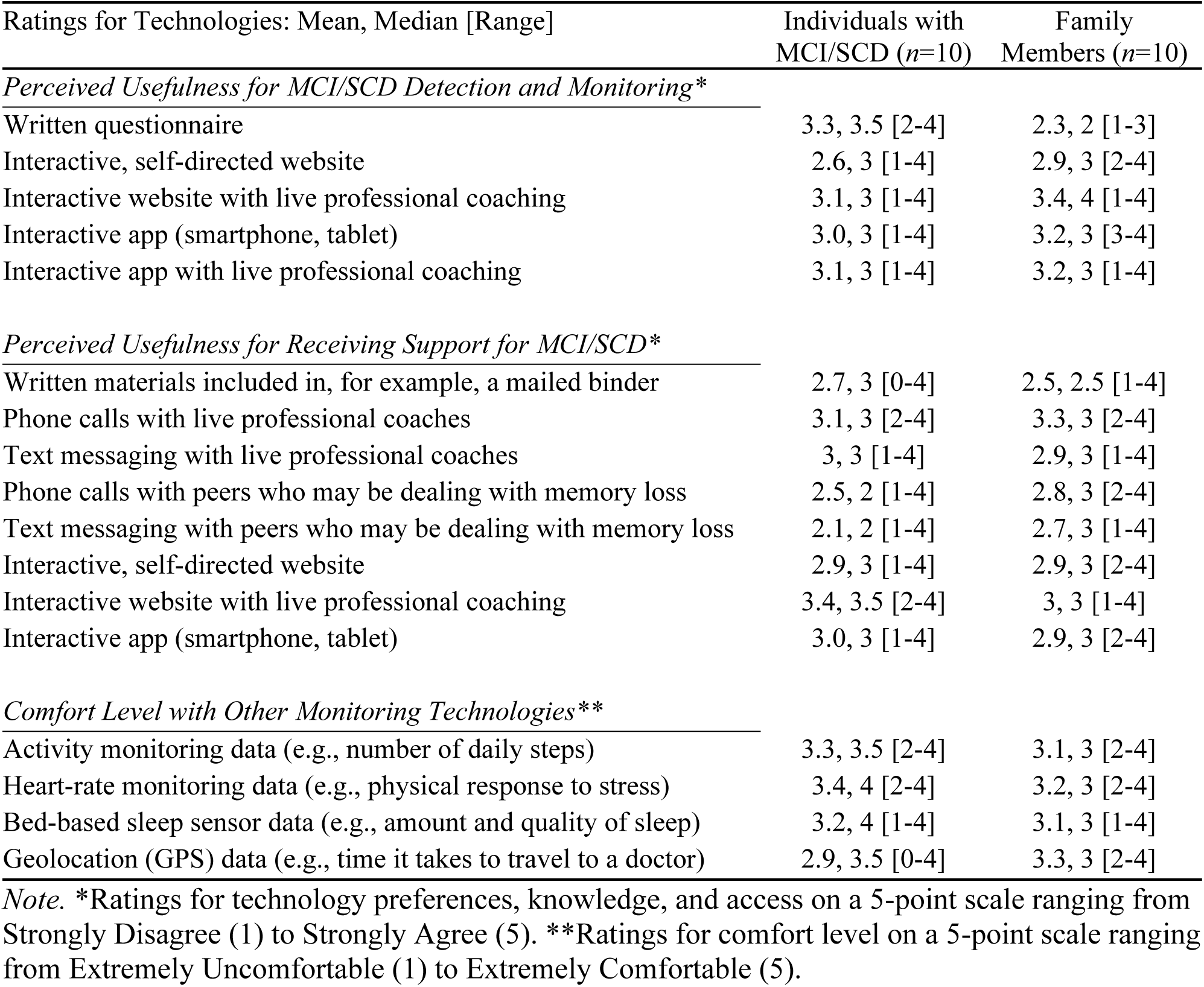
Participant Technology Preferences.

### Qualitative interview findings

Qualitative analyses suggested that participants were generally comfortable with the idea of using technology for MCI/SCD detection, monitoring, and support. However, they also had concerns. Factors influencing participant comfort with technology included sociodemographic characteristics, contextual factors, awareness of technology and perceptions about potential usefulness, actual technology use, and positive views of technology specifically as a tool for promoting the safety and well-being of the person with MCI/SCD.

For example, individuals with a background or career in technology-related industries, or access to friends or family members with such a background, expressed comfort with technology: *“I’m a millwright by trade… As foreman and superintendent, I had to do all kinds of tracking of time and for payroll and things like that. Ordering of materials and so forth*.

*Computers are not a problem… I’m fairly familiar with what’s going on.”* Participants also mentioned the availability of classes or other forms of training and support as contextual factors impacting their level of comfort with technology: *“… I would have to have somebody patient that would work with me so that I was on my own to do it and so that I could do it over and over again… I would have to have somebody teach me.”*

Participant awareness of and perceptions about the potential for technology to be useful had a bearing on the extent to which they would be comfortable using it for MCI/SCD detection, monitoring, and support. One individual commented, *“I know there’s different… apps and things like that, that kind of help with tasks and timers. I am really big on advocating for brain exercising and then trying to utilize some of those things. So, I would say that those things [would] definitely help me…”* Participants who used technology currently were also more comfortable with its use in the context of MCI/SCD. In describing use of technology for communicating with healthcare providers, one participant stated, *“We didn’t have to make an appointment. I didn’t have to drive anywhere, I didn’t have to park, I didn’t have to figure out where I’m going… It’s efficiency, really a lot of efficiency… I think people are more apt to communicate more when it’s easier.”* When commenting on the potential of technology for helping ensure the safety of the person with MCI/SCD, one family member commented, *“If you had an app where I could find out what was going on a little bit more and to be able to call her and prod her into action, that would be amazing. I would love that. Please make that.”* Concerns about technology were driven by sociodemographic characteristics, financial costs, fears about privacy and security, and the potential dehumanizing nature of technology.

Participants expressed “generational” challenges with embracing technology. One participant remarked, *“I think we are dealing with a generation… who didn’t learn to use these tools and in some ways, looks at them as being a distraction or a takeaway…”.* Another observed, *“…a huge part of … the medical field has been tablets and things… we’re just handing computers to people that have no idea how to do any of that…”.* Regarding her grandmother, another participant related how, *“…when [her] grandmother got older, she trusted technology less and less.”*

The financial aspect of using technology for MCI/SCD detection, monitoring, and support was another significant and recurring theme. One participant explained, *“The main reason why I don’t have hot spots is because I don’t want to pay for it.”* When describing barriers to using technology, another stated, *“At this current time, no, I don’t see any hindrances except finances…”*, and another asked rhetorically, *“What are the emerging technologies that are affordable to older people?”*

Privacy and security were also significant and recurring concerns. One participant remarked, *“They get into your computer and get your information…”, and another stated, “I worry about being hacked, giving out too much personal information.”* A different participant described how, *“… the big brother thing really does scare me.”* Others described how, in an increasingly technological world, *“… [they] feel anxious about sharing things like [they]were sharing before” and that, in some cases they feel that, “…the entire purpose of the app is to collect your data and drive you to using the app”*.

Finally, participants expressed concerns about what they perceived as the dehumanizing nature of technology. One person stated, *“I think that we depend too much on it and that we need to have more one-on-one interaction.”* Another explained how, in their view, *“Sometimes just having a person in front of you opens up the question-answer process… Sometimes just looking at a screen, it’s … Well, you’re really not there. I’m just looking at photons coming from the computer screen, and that kind of is a problem.”*

## Discussion

This study explored access, knowledge, and technology preferences among rural individuals with MCI/SCD and their family members to gain insight into how best to monitor and support cognitive health in these communities. An effective strategy for developing supportive technologies may be to leverage capabilities that rural families value (e.g., to organize and track tasks, to “exercise” one’s brain, to communicate with healthcare providers, to promote safety of the person with MCI/SCD) while simultaneously addressing the concerns that they have.

One such concern is that older rural adults may not be interested in, or able to, engage with digital technology. This is a misconception that runs counter to data showing that technology use has increased dramatically among older adults in recent decades (e.g., 13% of adults aged 65+ owned a smartphone in 2010 versus 61% in 2021; 6% of adults aged 65+ owned a tablet in 2012 versus 44% in 2021; (34)) and that mobile devices have continued to proliferate in rural areas (35), with as many as 87% of rural adults reporting that they use a smartphone (36). Education and awareness about these trends, as well as efforts to increase digital health literacy, may boost confidence in the potential of digital health approaches for supporting cognitive health among rural families which will ultimately lead to improved outcomes (37).

Another concern has to do with cost. In this study, interview participants reported that the cost of technology was a significant barrier to adoption and use, which is consistent with existing research (24). However, in the context of the present sample in which 55% of participants indicated being financially “comfortable” (i.e., as opposed to “just enough…” or “not having enough to pay for living expenses”), it is possible that some individuals are not aware of the degree to which devices such as smart phones and tablets have grown more affordable in recent years. Then again, the cost of devices themselves may be only a piece of the equation for rural families with limited and fixed resources who must often prioritize other basic necessities such as housing, utilities, prescriptions, and food.

Educating older adults who have adequate financial resources about the technology required for MCI/SCD detection, monitoring, and support (i.e., that simple and inexpensive devices may be sufficient) is one strategy to allay concerns about costs. A second strategy is to engage with policy makers to reframe access to these technologies as a basic necessity, comparable to those previously mentioned. From a policy perspective, it is noteworthy that federal efforts to increase and sustain access to digital technology for older adults and those living in rural areas (e.g., Digital Equity Act of 2021) are currently under threat. It is urgent that developers of digital supports for rural families contending with MCI/SCD work with policymakers to advocate for continued funding of critical programs (38).

A third concern that must be addressed involves security and data privacy. As with cost concerns, these findings about security and data privacy were nuanced with qualitative interview data showing that participants were concerned about security and data privacy but survey data showing that they were open to the idea of using other types of data to better understand families’ experiences with MCI/SCD (e.g., daily step counts, physical stress response, amount and quality of sleep, time it takes to travel to a doctor). These apparent contradictions demonstrate the complexity of these issues, where families may be leery of, but also intrigued by, technology. Similar to overcoming concerns about age and cost as barriers to technology use, specific, transparent, and accessible training may be effective for educating users about how data are secured and user privacy is protected. This is important in light of research demonstrating the potential of consumer technologies such as smartphone-based GPS to detect and monitor cognitive decline (39). Our findings also highlight that a one-size-fits-all approach to training and implementation may not be the best since individuals have different levels of comfort with technology based upon their education, experience, and career history.

In terms of preferences for detection and monitoring of MCI/SCD, survey data evidenced discrepant opinions between individuals with MCI/SCD and their family members, with the former preferring simple written questionnaires and the latter preferring interactive websites with live professional coaching. However, for receiving support for MCI/SCD, there was general consensus that interactive websites with live professional coaching would be an effective strategy. This was reinforced by qualitative data about the risks of technology eclipsing human interactions.

Surprisingly, peer-support (e.g., phone calls or text messages from peers) was not highly preferred, particularly by individuals with MCI/SCD, despite literature documenting this as a promising approach for health interventions broadly (40) and memory programs specifically (41). What may be at issue here, is the meaning of the term “peer” and whether hesitancy about this approach could be overcome by systematic training of peer supporters. An additional consideration may be internalized stigma associated with memory loss (42), as well as fear of stigmatization by peers. These are important questions considering the potential of senior centers to provide programmatic support to meet needs in under-resourced, underserved, and isolated rural communities (43).

Although this study sheds light on technology access, knowledge, and preferences of rural individuals with MCI/SCD and their family members for supporting cognitive health, at least two limitations must be noted. First, the convenience sample was predominantly female, white, and financially comfortable. One hundred percent of participants in this study indicated having access to and being knowledgeable about the Internet/Wi-Fi and, with the exception of iPads and iPhones, they overwhelmingly had access to other technologies for potential detection, monitoring, and support of MCI/SCD.

These findings could be partly attributable to the strategy of partnering with senior centers for participant recruitment. Senior centers may attract a unique subpopulation of active and “connected” older adults who are engaged with their community and have the interest and capacity to participate in research. Moreover, although the authors set out to understand the experiences of MCI/SCD in rural communities, many of the sample resided in what may be considered “small towns” which are generally better connected. However, the lack of availability of memory assessment and care in these communities mirrors that of more geographically isolated communities. Expanding recruitment strategies (e.g., working with healthcare and culturally specific social service providers for study referrals) could yield a sample with a greater diversity of experiences with technology and, thus, increase the applicability of findings. This expanded recruitment in future research may provide additional insights into differences in access to these technologies, as well as begin to distinguish *access to* vs. *actual use of* these technologies.

Second, although the survey data provided valuable information, our sample was small and limited to providing descriptive contextual information for the qualitative interviews. A larger sample of survey participants would yield more robust data and provide for greater confidence in our conclusions about technology access, knowledge, and preferences among rural families experiencing MCI/SCD. Further, due to our small sample size, we grouped together participants with MCI and/or SCD. An important next step for future research is to evaluate the potential for unique perspectives to emerge between participants with MCI versus participants with SCD, as well as across other cognitive statuses (e.g., cognitively unimpaired, MCI, SCD, Alzheimer’s disease and related dementias).

## Conclusion

High rates of MCI/SCD (1,2) in the United States, as well as the potential for these conditions to develop into Alzheimer’s Disease (9), necessitate that individuals with MCI/SCD and their family members be provided with supportive resources, regardless of where they live. Rural life presents a variety of challenges for detection, monitoring, and support of brain health (10–14). However, these challenges may be creatively addressed through user-friendly technologies delivered in community settings such as Senior Centers. This study provides valuable information with which to pursue this goal and, ultimately, improve cognitive health in rural communities.

## Data Availability

Data for this study are available from the corresponding author upon request.

## Acknowledgments

Funding for this study was provided by the Arizona Alzheimer’s Consortium: https://azalz.org/ (recipients MJM, MM, ESC). The funders had no role in study design, data collection and analysis, decision to publish, or preparation of the manuscript. We also wish to thank our Senior Center partners in Flagstaff, Bullhead City, Prescott, Prescott Valley, Snowflake, and Yarnell, AZ, with whom we have ongoing collaborations for participant recruitment, data collection and dissemination, and co-creation of brain health supports.

